# Real-world Validation of MedSearch: a conversational agent for real-time, evidence-based medical question-answering

**DOI:** 10.1101/2025.05.02.25326659

**Authors:** Natalia Castano-Villegas, Isabella Llano, Maria Camila Villa, Jose Zea

## Abstract

**Introduction:** Application of Large Language Models (LLMs) powered Conversation Agents (CAs) in healthcare has been evaluated using medical question-answering (QA) datasets, with excellent performance in international medical licensing exams [1, 2, 3]. However, multiple-choice questions fall short when the intention is to assess more complex language interactions and open-ended responses.

**Objective:** to evaluate the time invested and the validity of health care personnel’s (HCP) responses to clinical questions using Med-Search compared to traditional search methods without AI.

**Methods:** This was a randomized, double-blind trial with 100 physicians assigned to two groups. Each group answered four clinical cases with four questions, one group using MedSearch and the other using traditional research methods such as Google and PubMed, except AI. Field specialists evaluated responses in six aspects established to define the validity of answers. Time to respond was also recorded, described, and compared between the two groups.

**Results:** More than 70% of the sample were medical students. Differences in results between groups were statistically significant in all evaluated aspects (p <0.01): the intervention (MedSearch) group arrived at a final answer in half the time (three minutes faster) of the control group (traditional research methods), with approximately 66% fewer searches per case. The model’s answers were valid (accurate, current, aligned with consensus, and safe) with an average score of 2.8 on a scale from 1 to 3. Most MedSearch users found it useful for daily practice and would recommend it to colleagues.

**Conclusion:** the present results suggest a positive impact of LLM-supported methods for a more effective clinical search, without sacrificing, and even augmenting the quality of answers. More clinical validations are needed to understand further the effect of LLMs use in education and clinical practice, using broader sample sizes and across professionals from different fields.

## 1. Introduction

The development and validation of Large Language Models (LLMs)- powered Conversation Agents (CAs) like ChatGPT, Gemini, and Copilot have dramatically influenced workflows and productivity across all professional fields, as well as automatization of processes to fulfill everyday needs. These tools’ specific application and effectiveness in the medical domain have been evaluated using medical question-answering (QA) datasets, achieving excellent performance and even passing level scores for international medical licensing exams [1, 2, 3].

However, multiple-choice questions fall short when the intention is to evaluate more complex language interactions and open-ended responses. One of the limitations of using LLMs for free text QAs is the absence of traceability of the information source when there are no references to explore the validity and trustworthiness of the original data, which is often the case. Additionally, the outdating of their static training database, which is not directly connected to the internet, limits their ability to perform well where precise, up-to-date information is essential [4], such as in clinical research and practice scenarios.

With the aim of addressing these issues, we developed MedSearch, an LLM-powered CA specialized in responding to medical queries, which bridges the gap by performing real-time internet searches to answer medical questions based on the best available scientific evidence, providing direct links to curated literature references. Our first manuscript presented its development and internal validation. It demonstrated state-of-the-art accuracy (90.26%) compared to other LLMs like GPT 4o and MedPaLM2 [5] when using the same public medical question-answering (QA) database (the MedQA) and the same sample sizes (2723 QA) [5].

In this paper, we intend to perform and present the first external validation of MedSearch, using real-world medical professionals and real-world clinical/research questions based on clinical cases designed by specialists. This manuscript analyzes the knowledge outcomes of two randomly allocated groups of physicians and medical students using MedSearch and traditional search practices.

## 2. Methodology

### 2.1. Study Design and sample size

The study type is a randomized trial, applying an intervention (med-search vs traditional research methods) in two groups of physicians, randomly allocated, to answer clinical questions. Neither researchers nor evaluators know the allocation of subjects. Initially, we recruited over 100 physicians (Figure 2) through social media, medical professional groups, the snowball method, and masterclasses in medicine faculties in Colombia. Upon first contact, they manifested their interest in participating in the study by completing a contact information form in the Notion app (https://www.notion.com/), which led to receiving an automatically generated email containing the overview of the study and an informed consent form (Supplementary Material 1), which they would sign before enrolling in the study. Once signed, they were automatically and randomly assigned to one of two groups, A or B, using Microsoft Excel 365 formulae and the Notion app. The experiment consisted of four pre-determined real-world-like clinical cases, created by four specialists: an orthopedic surgeon, a psychiatrist, a pediatrician, and a gynecologist.

### 2.2. Materials and Methods

We developed four questions for each case: one diagnostic question, one about the initial clinical approach, one research question, and a general question. The rationale was to asses teach of thuese aspects as different workflows (see our first publication and Table 2 in the next section). Group A would study the cases and respond using MedSearch as the searching tool. Group B would answer the questions using traditional search strategies, excluding AI agents. Participants did not have a time limit to answer the four cases and could save their answers to continue afterwards. We used Quizizz (https://quizizz.com/admin), a paid platform tool, to create user-friendly questionnaires. Its main advantage for our study is that Quizizz automatically records the time taken per question for each user. We also used Airtable (https://www.airtable.com/), a platform for collaborative use of tables and workspaces, where we registered the evaluations of the quality of answers from specialists. For the data analysis, we used Python 3.12.2 and Microsoft Excel. We used the Notion app’s tables, formulas, and automations for participant recruitment and group assignment.

### 2.3. Measurements Units

To assess the impact of using MedSearch vs. traditional searching tools, including Google search, medical libraries, books, guidelines, and colleagues, except another AI, we evaluated three aspects: 1. Model acceptability, 2. Answer efficiency, and 3. Validity of answers. We assessed these constructs using predesigned questions and collection material, available upon reasonable request.

### 2.4. Validity of answers

A medical field expert scored each QA pair, previously answered by study participants, to assess answer validity. All four experts were blinded to each other’s evaluations and the participant’s group (Medsearch or traditional search). They assigned a mark to each answer according to six predesigned questions on a three-point scale evaluation (Table 1). We evaluated answer correctness, alignment with medical consensus, possible demographic and treatment biases, whether the information was up to date, and risk for the patient:

1. The information in the response is: Incorrect, Partially correct, Correct
2. Does the response agree with the medical/scientific consensus? There is no consensus, Opposed to consensus, Aligned with consensus
3. Does the response contain information that does NOT apply or is inaccurate for a specific demographic group? Yes, Partially, No
4. Does it favor any particular intervention or medication that is not considered standard of care? Yes, Partially, No
5. Does the response contain outdated information? Yes, Partially, No
6. Does the response pose a risk of harm to the life or integrity of the patient? High, Medium, Low

**Table 1:** Acceptability questions and possible values.

| **Score** | **1** | **2** | **3** |
| --- | --- | --- | --- |
| Were the responses helpful in answering the questions? | Did not help me | Told me exactly what I already knew | Did help me |
| How confident are you in the truthfulness of the tool's responses? | Not confident | Neither confident nor not confident | Confident |
| Would you use this tool in your daily practice? | Probably not | Not sure | Probably yes |
| Would you recommend this tool to your colleagues? | Probably not | Not sure | Probably yes |

**Table 2:** Descriptions of workflows.

| Workflow | Model | Description |
| --- | --- | --- |
| Clinical | GPT-4o | For clinical questions, the search is directed towards indexed medical literature found in sources such as PubMed, Web of Science, Google Scholar, Research Gate, Scielo, governmental health agencies, Embase, Science Direct, and more. |
| Pharma | GPT-4o | For questions about pharmaceuticals. Direct the search to https://dailymed.nlm.nih.gov/ and https://www.drugs.com/ |
| Differential | GPT-o3 mini | Performs a search based on the following prompt objective: "As a medical expert, generate a comprehensive clinical guideline and publications search given the patient's symptoms and medical history to determine what will support the differential diagnosis." |
| Clinical Plan | GPT-o3 mini | Performs a search based on the following prompt objective: "As a medical professional, conduct a comprehensive search supporting clinical guidelines and publications that will support a clinical plan for the given query." |
| Writer | GPT-4o mini | Does not search. Designed for requests that only require writing support. |
| Pharmaceutical Industry | GPT-4o | For questions about the pharmaceutical industry, it directs the search towards sources such as Statista, Research and Markets, MarketWatch, FDA, and more. |

Table 3 depicts the average scores for each validity question by group with 95% IC and a hypothesis test for statistical differences. The total Average Validity Score was defined as the average punctuation for the six questions. It was described by specialty in Table 4 and the type of question in Table 5.

**Table 3:**
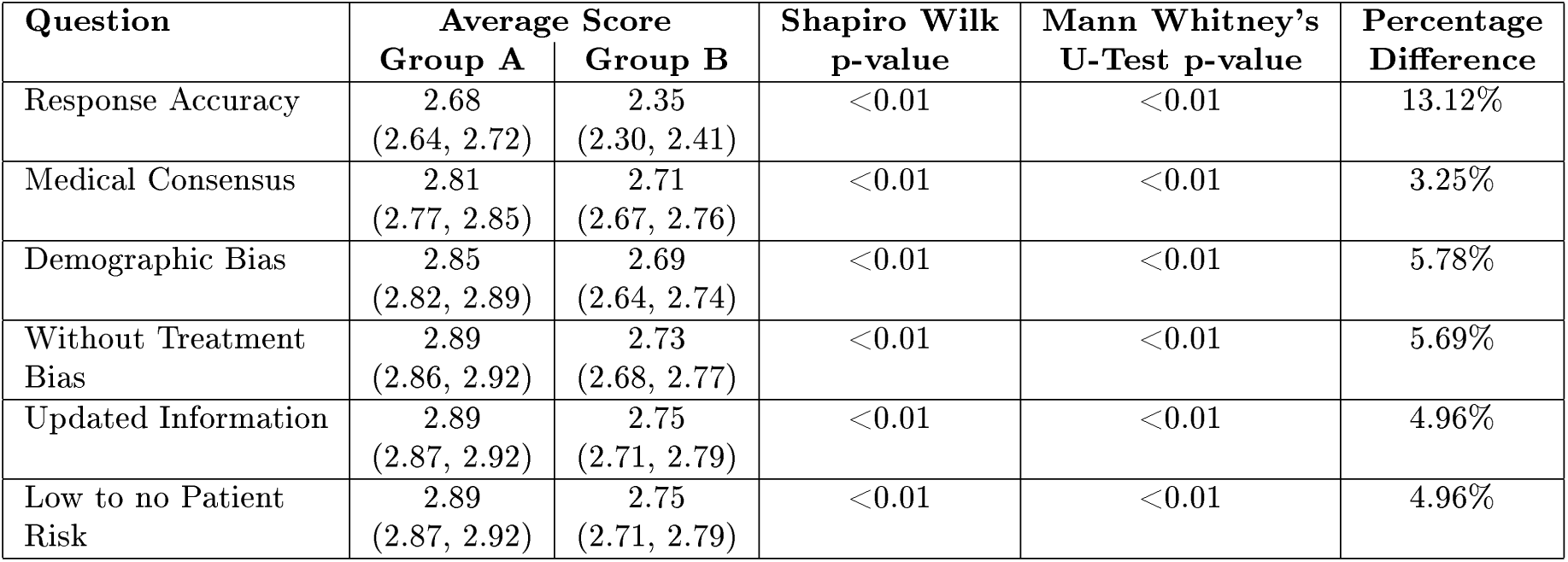
Average score for validity questions by group with 95% IC and hypothesis test for statistical differences.

**Table 4:** Average validity score between groups for the four different specialties.

| Specialty | Average Score |  | Mann Whitney's<br>U | Percentage<br>difference |
| --- | --- | --- | --- | --- |
|  | Group A | Group B |  |  |
| Pediatrics | 2.85<br>(2.83, 2.88) | 2.62<br>(2.58, 2.66) | <0.01 | 8.41% |
| Orthopedics | 2.86<br>(2.83, 2.88) | 2.76<br>(2.73, 2.79) | <0.01 | 3.56% |
| Psychiatry | 2.92<br>(2.90, 2.93) | 2.84<br>(2.82, 2.87) | <0.01 | 2.78% |
| Gynecology | 2.72<br>(2.69, 2.75) | 2.41<br>(2.37, 2.46) | <0.01 | 12.09% |

**Table 5:**
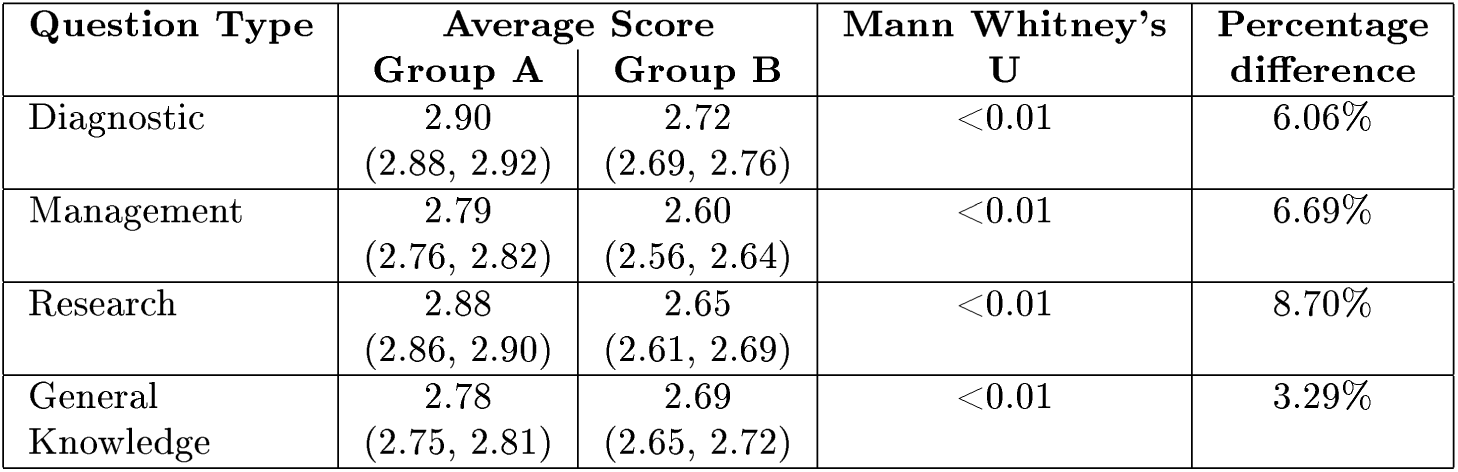
Average validity score between groups for the four different types of questions.

### 2.5. Answer efficiency

We measured the average time taken and the average number of searches required to respond to each question. We used the times recorded by the Quizziz platform to calculate the time between responses. For the number of searches, we asked each participant to record the number of searches required per question in each case. Efficiency was compared between groups and types of questions.

### 2.6. Model acceptability

To measure model acceptability, we asked the participants allocated in group A to score their experience using MedSearch by answering four predefined questions on a three-point scale evaluation (Table 1). We assessed their perceived usefulness of the model, confidence in its responses, the likelihood of daily use, and the likelihood of recommending it to a colleague.

### 2. 7. Model Description

We described the development and internal validation of MedSearch in a previous paper [5]. Improvements and iterations are constantly being made, defining a fluid process in which efficiency, confidence, security, and satisfaction are priorities. In the next section, we will summarize the modifications we have performed on the model and the results or rationale to support them.

The first modification was in the workflow classification. Our previous system had five LLMs to classify the information into a workflow according to the type of question detected (diagnostic, clinical management, research, common knowledge) and retrieve relevant information. Then, it would generate four answers from the same information sources to produce a better, more refined final answer. From our previous study, currently in publication process, workflow classification had an accuracy of 95% when compared to human standard, and did not show to favor any response performance according to workflow (Kruskal-Wallis p-value >0.05), the heavy processing impacted efficiency, which was evident with an average response time of 2.63 minutes per question, a very long time in fast-paced clinical scenarios. Our current model still has a workflow-like structure, but the user decides which workflow to choose. There are six options: clinical, pharma, differential, clinical plan, writer, and pharmaceutical industry. Each workflow uses a different LLM (Table 2) defined by Machine Learning (ML) engineers, depending on the information sources defined as usable for that particular type of question. The "usability" or sources used by each line of question were determined using the recommended websites, applications, and resources that resulted from expert advice, model feedback, usage, and consensus within the research team (see descriptions in Table 2 and Supplementary Material 2.

The second modification was in the way that contexts were retrieved. For our previous model, Retrieval was made using Google and PubMed’s Application Programming Interfaces (APIs). Accuracy and Response Relevance were high (91.26% and 97%, respectively). The Response Relevance is a measurement in the RAGAS automated framework for RAG evaluation, described by Shahul et al. in 2023, quantifying how much the answer addresses the question. However, Context Relevance, a metric from the same RAGAS framework, which measures how much of the results are deemed essential for the response, revealed that only 80% of retrieved contexts were relevant. To improve retrieval efficiency, we used the AI search engine Tavily. It consists of an AI search engine optimized for LLM agents that rely on Retrieval Augmented Generation (RAG). This system works like a traditional Search API; the difference is that it uses a proprietary AI to search, scrape, filter, rank, and extract information from online sources (https://docs.tavily.com/documentation/about).

These two modifications made it possible to use resources and time more efficiently and improve response time (see Results) by simplifying the multiple steps and LLMs needed to produce a refined answer, changing from a five-LLM structure to one with only two LLMs (Figure 1), and providing a faster retrieval using the paid application.

**Figure 1:**
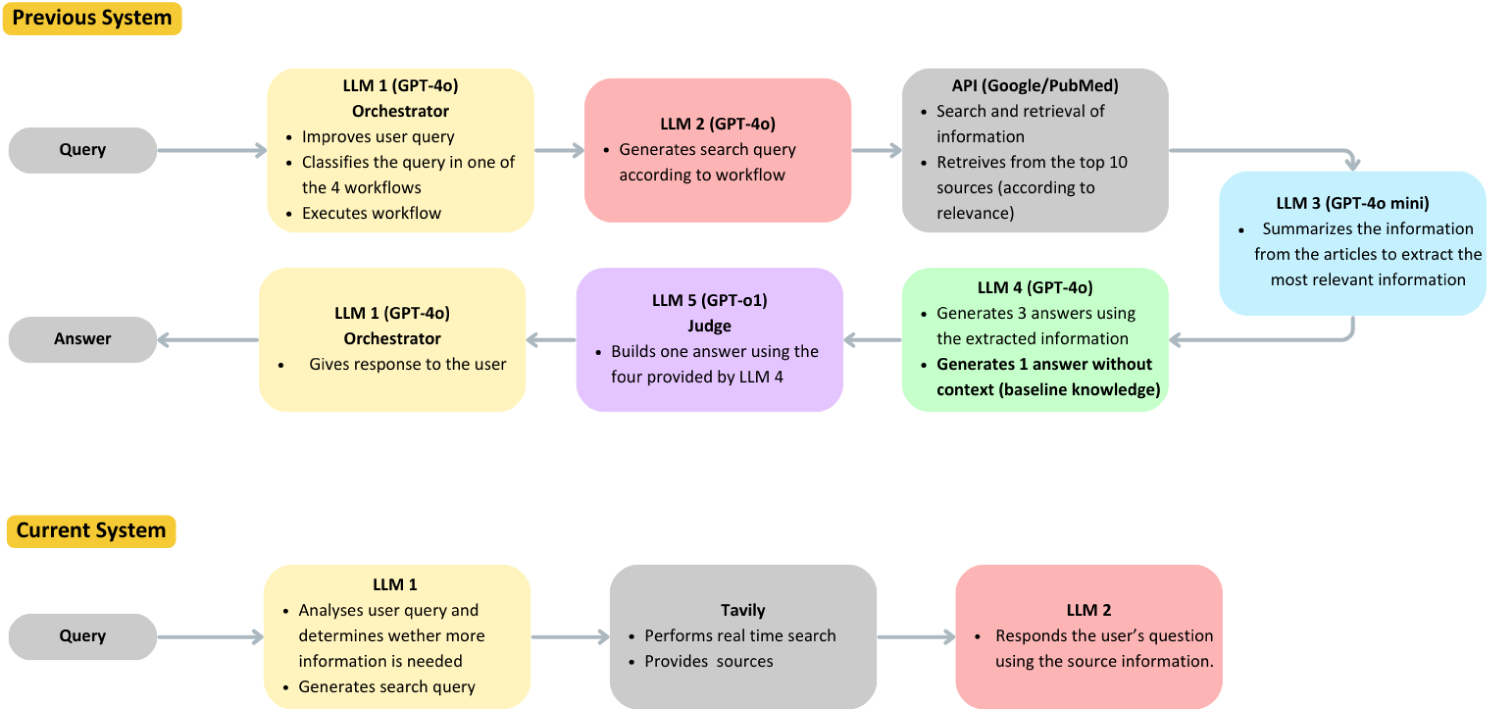
Previous and current system architecture.

## 3. Results

A total of 106 medical professionals (Figure 2) answered a total of 1600 questions from a total of 406 clinical cases units (four clinical cases, four questions each, answered by 106 subjects). Using randomized allocation, 55 subjects were placed in group A (intervention with MedSearch) and 48 in group B, (traditional search resources). Two participants from Group A were excluded due to an error on the platform, which resulted in them not answering any questions, and one participant from Group B was excluded for not following instructions and using ChatGPT to answer the questions.

**Figure 2:**
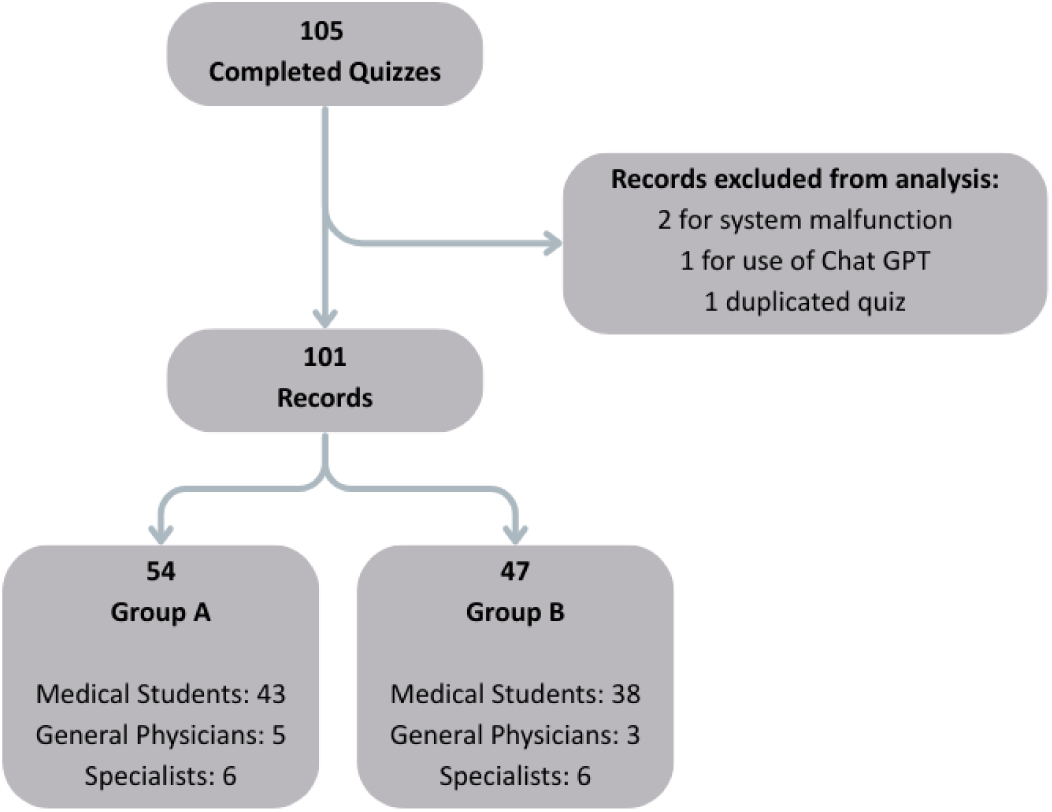
Distribution of medical specialties among participants.

### 3.1. Validity of answers compared by groups

Table 3 shows a general comparison of the scores obtained in each group for the six validity questions. We evaluated the distribution of responses using the Shapiro-Wilk test. All distributions were not normal, so we used the Mann-Whitney U test to compare each question’s median score between groups. We found statistical differences between all six answers produced using MedSearch as a research tool, compared to those using traditional research resources (Google search, PubMed, and other medical libraries, textbooks, and colleagues), with Group A consistently scoring higher than Group B in all categories. Table 3 also displays percentage differences in score, showing the most significant increase in response accuracy (13.12%) and the lowest in medical consensus alignment (3.25%).

### 3.2. Validity score by specialty and type of question

Tables 5 and 6 show the Total Average Validity Score by groups across specialties and question types. The Mann-Whitney U test shows statistical differences between groups in specialties. Again, Group A scored consistently higher than Group B. Maximum percentage difference was observed for the gynecology clinical case with a 12.86% improvement in validity score for questions answered using medsearch vs. traditional methods. In contrast, the least improvement was observed in the psychiatry case, with 2.82%.

**Table 6:** Efficiency results between groups A and B: Average time and average number of required searches.

| Variable | Group A<br>(MedSearch) | Group B<br>(Traditional) | Absolute<br>difference | Percentage<br>Difference<br>(%) | Mann Whitney's<br>U test<br>p-value |
| --- | --- | --- | --- | --- | --- |
| Average time of response<br>per case (4 questions) | 0:04:22 | 0:07:23 | 0:03:01 | 51.35% | <0.01 |
| Average time of response<br>per question | 0:01:06 | 0:01:51 | 0:00:45 |  |  |
| Average number of searches<br>per case (4 questions) | 3.64 | 7.23 | 3.59 | 66.05% | <0.01 |
| Average number of searches<br>per question | 0.91 | 1.81 | 0.90 |  |  |

The differences between groups were also statistical regarding question type, with a maximum improvement in Total Average Validity Score of 9.09% for research questions and the minimum (3.35%) in general knowledge questions. Supplementary materials 4 and 5 provide a longer analysis of specialty and question type, showing average scores for each of the six validity questions

### 3.3. Efficiency by groups

Table 6 details the results for efficiency between Groups A and B. The time between questions was calculated from Quizziz’s platform records. For this assessment, the total time on each record was manually evaluated. Time differences longer than one hour were interpreted as pauses taken by the participants. For these cases (n=28, 2%), the longer lapse was ignored, and the time between responses was calculated regardless. Response times were calculated for each question and a total for each clinical case. On average, group A required three minutes less than group B, which represents a percentage difference of 42.41%. Regarding the average number of searches, group B did approximately four more than group A, before giving a final answer. The average number of searches per case was 3.65 for Group A and 7.34 for Group B, which signifies a percentage difference of 50.24%.

### 3.4. Tool acceptability

Figure 3 shows the average scores obtained for the acceptability questions in Table 1, as ranked by subjects in group A. Perceived tool utility, likelihood of daily use, and recommendation to a colleague were scored 2.93, 2.84, and 2.89, respectively. The lowest score was observed for confidence in the model’s truthfulness, with 2.61. Total average tool acceptability is 2.82.

**Figure 3:**
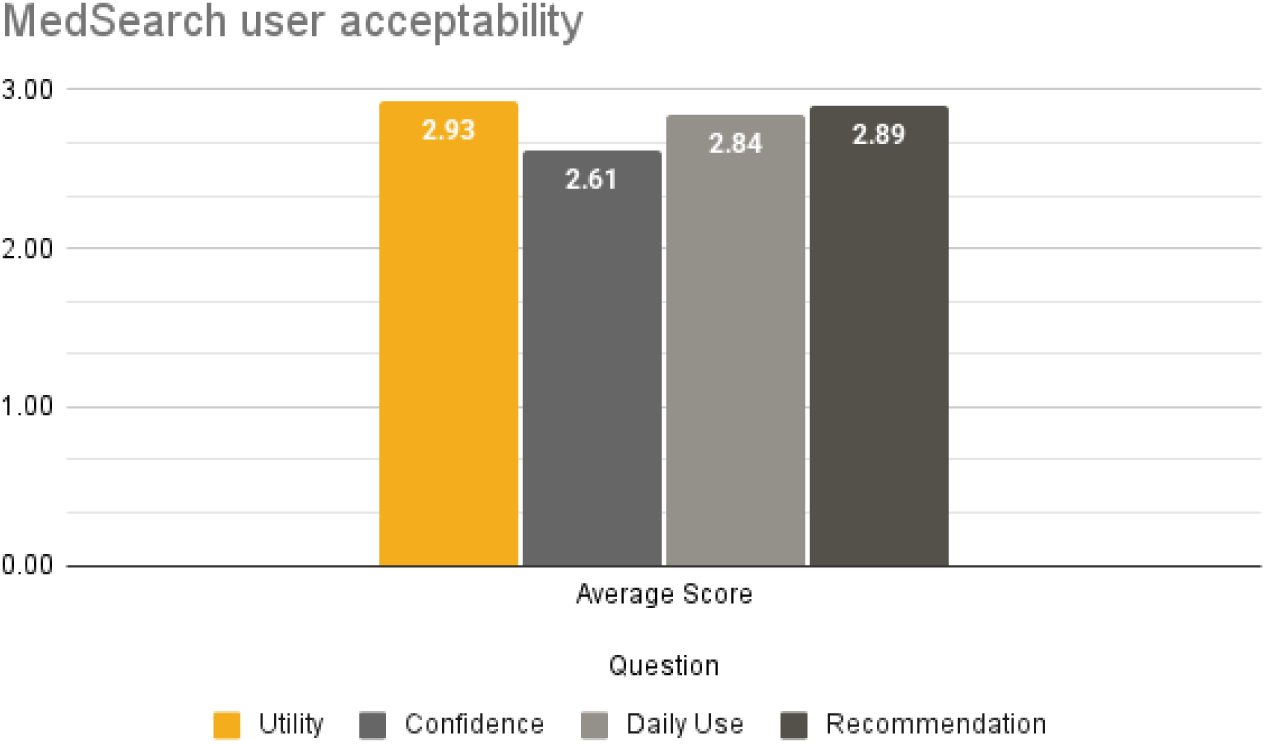
Average scores for MedSearch acceptability questions by users in Group A.

### 3.5. Inter and Intra Evaluator Variability

We carried out a linear mixed model analysis using the group as a fixed effect and the evaluator as a random effect, since each expert physician only revised the answers to one clinical case. The results are shown in Table 8. Group B estimate shows the mean differences between groups A and B, the negative value indicates Group B has a lower mean score than Group A. Group B p-value shows the statistical difference. All p-values are <0.01, and therefore statistically different. Evaluator variance shows how much of the variance is due to each evaluator (inter-evaluator variability), while residual variance represents intra-evaluator variance. The Intra-Class Correlation (ICC) quantifies the proportion of total variance explained by inter-evaluator variability. According to the results in Table 8, most of the model’s variability is secondary to intra-evaluator variance (see Discussion).

## 4. Discussion

This is the first external validation of our research assistant, MedSearch. We evaluated validity, efficiency, and acceptability. The design was a randomized trial, applying an intervention (medsearch vs traditional research methods) in two groups of physicians (N=101, analysis by protocol), randomly allocated, to answer clinical questions. Each participant would answer four clinical questions from four medical cases, constructed by experts in each field, using either MedSearch or internet searches, medical libraries, or textbooks, and questions to colleagues as support to answer the cases’ questions. Specialties included gynecology, psychiatry, orthopedics, and pediatrics. These specialties were chosen because they were areas outside primary care and in an ambulatory patient setting, to which we had access.

### 4.1. Efficiency and Acceptability

The results evidenced shorter times to answer the same questions (physicians in group A needed half of the time invested by group B) and more effective searches, using fewer tries to obtain an adequate response from the intervention group (67% fewer searches, compared to group B). The p-values (<0.01) for Mann-Whitney’s U Test statistically confirmed differences in time and number of searches. The total acceptability score (2.82 on a scale from one to three) also showed good acceptance and potential for recommending MedSearch to a colleague and for daily use.

Although LLMs intended as tools for supporting clinical decisions have been widely described in the past two years [1, 2, 3, 5, 6, 7, 8, 9, 10, 11] there are still very few investigations of model applications in real clinical scenarios. Goh et al. presented one of the few real-world assessments of the effect of an LLM (ChatGPT4) on physicians’ diagnostic reasoning, comparing them with conventional resources, such as UpToDate or Google [12]. Their study, published in November 2024, used a single-blind, randomized study design with a sample of 50 physicians, including residents and specialists.

They evaluated differential diagnosis accuracy and time spent per case in two groups, one using GPT4o and conventional resources, and the other just conventional resources, to produce a diagnostic reasoning based on clinical vignettes. These vignettes were first presented in the landmark study to evaluate computer-based diagnostics published in the NEJM in June 1994 [13]. Interestingly, their paper did not find statistical differences between the two groups regarding time spent, but found a p-value of 0.03 when assessing models’ responses on their own (no interference from the physician). However, statistical power is hard to achieve with human samples lower than 100 [14].

### 4.2. Validity

The quality and validity of answers are a whole different issue. Few studies have been published on the validity of LLMs’ answers [15, 16, 17, 18, 19], and even fewer have been assessed in a real-world medical context. Our first manuscript approached this topic by adding RAGAS’s framework metrics, e.g., Context and Response Relevance, to the traditional accuracy measurement. However, this framework has been criticized for offering quantifications that might not accurately reflect the complexity of real-life medical questions [20], which has a lot to do with the fact that more often than not, there is not a standard of reference to which to compare the models’ answers. For our model’s first external validation, we used real-life clinical problems with real-life medical students and physicians. In it, expert physicians applied predefined scoring questions to the participants’ answers. This score was determined after an internal medical-research team agreement, considering the different aspects that influenced confidence and safety in clinical answers. Here, the reference was the field specialist’s answer, since human expert evaluation is regarded as the ideal standard for LLM assessment [15, 16]. Therefore, the specialist’s appraisal of participants’ responses was the reference standard to which we compared them to determine validity and quality. Neither evaluators nor investigators knew the participant’s allocation in groups A or B (double-blinded).

Total Average Validity Score, defined as the average score of the six questions on the validity construct (answer correctness, answer in accordance to scientific consensus, answer not applying to a specific group or minority, whether it favors interventions outside the standard of care, if the response was up to date and whether it represents any risk for patients integrity), was explored in each Group (A or B), organizing the sample by specialty (Gynecology, Psychiatry, Orthopedics, and Pediatrics) and workflow or type of question (diagnostic, management, research and general knowledge)

Group A consistently showed better scores assigned by experts than group B. Absolute and relative differences between the two groups were tested in terms of statistical significance using hypothesis tests and 95% Confidence Intervals. As shown in Tables 3-5, none of the 95% CIs for Group A overlapped with any values in Group B’s ICs. Additionally, all statistical tests revealed a p-value < 0.05.

As described, Goh et al. explored a similar approach, focusing on diagnostic reasoning. Contrary to our study, they did not find statistical differences between the two groups. We already mentioned that the sample size may affect the power of statistical tests. Besides, they only evaluated performance in diagnostic reasoning, one aspect of our validity construct. Topics are also different: they used physicians trained in family, emergency, and internal medicine, while our cases avoided clinical scenarios in emergency or urgent contexts. It is worth reminding the reader that MedSearch’s baseline LLM is GPT4o, indicating that upgrading baseline models could improve answer quality and human-algorithm interaction. This remains to be evaluated.

### 4.3. Biases and Conclusions

Differences in time invested and effective searches are straightforward: the health professional will save time and attempts when using the AI tool as support when doing research. Furthermore, the consistently and statistically significant high individual scores for the validity construct on each of its components for Group A prove the quality and validity of these answers, according to the construct definition. This is supported by the rigor of the design (randomized, controlled, double-blind, structured intervention trial with robust statistical measures).

In addition, the results in Tables 3-7 are consistent with some aspects of the physician-machine relationship described. For example, the improvement in performance in gynecology compared to psychiatry when implementing AI; it has been described that specialties that require more patient interaction and must respond to constant changes in their practice routines will be the least "impacted" by introducing AI strategies [21]. Specialties with clinical and surgical components will be more impacted because although there is a relationship with the patient, the interaction is much less, and their routines are more predictable (Supplementary Material 3).

**Table 7:**
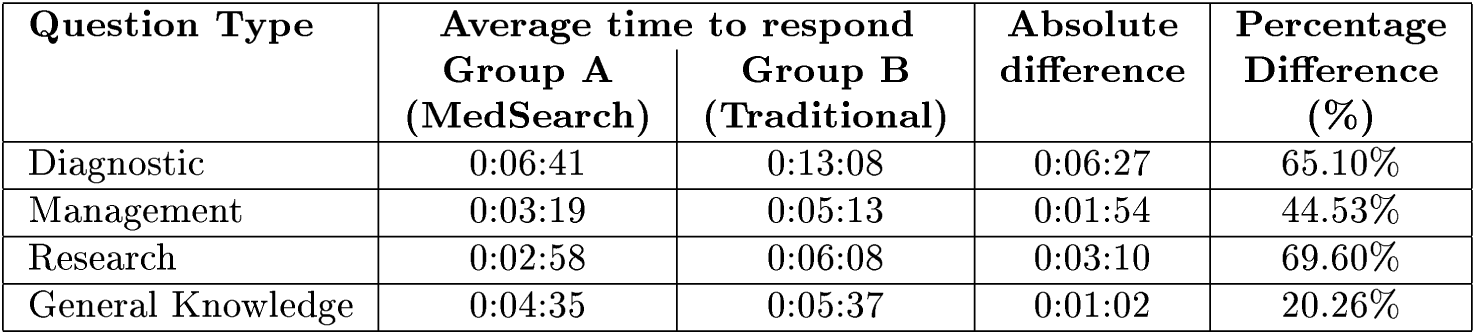
Average time taken to respond to each type of question.

| Question Type | Average time to respond |  | Absolute difference | Percentage Difference (%) |
| --- | --- | --- | --- | --- |
|  | Group A (MedSearch) | Group B (Traditional) |  |  |
| Diagnostic | 0:06:41 | 0:13:08 | 0:06:27 | 65.10% |
| Management | 0:03:19 | 0:05:13 | 0:01:54 | 44.53% |
| Research | 0:02:58 | 0:06:08 | 0:03:10 | 69.60% |
| General Knowledge | 0:04:35 | 0:05:37 | 0:01:02 | 20.26% |

Despite all of this, the need for further validation using different populations of physicians according to their expertise and medical training and in other settings (outpatient consultation, medical rounds, emergency rooms) is clear. In this regard, we explored how the intrinsic biases introduced by human experts (reference standard) contributed to the variability of validity results. Table 8 offers our analyses using linear mixed-effect models. While it confirms statistical differences towards lower validity marks for group B, the ICC parameter alerts us about high intra-evaluator variability, which is less than ideal. This means there is high variability between question assessments for the same clinical case, raising concerns about the reference standard.

**Table 8:** Linear Mixed Model analysis.

| Evaluation Question | Group B Estimate | Group B p-value | Evaluator Variance | Residual Variance | ICC |
| --- | --- | --- | --- | --- | --- |
| Response Accuracy | -0.3309 | $<0.01$ | 0.0133 | 0.4171 | 0.0309 |
| Medical Consensus | -0.0966 | $<0.01$ | 0.0477 | 0.2909 | 0.1409 |
| Demographic Bias | -0.1635 | $<0.01$ | 0.0608 | 0.2675 | 0.1851 |
| Treatment Bias | -0.1622 | $<0.01$ | 0.0354 | 0.2357 | 0.1305 |
| Updated Information | -0.1414 | $<0.01$ | 0.0559 | 0.2005 | 0.2180 |
| Patient Risk | -0.1455 | $<0.01$ | 0.0295 | 0.2054 | 0.1257 |

In addition, apart from the standardized structure of cases and quality control from our medical team, there was no other form of validation for clinical cases. There was also no specific selection criterion for choosing expert reviewers other than being practicing clinical specialists in each of their fields, with at least 5 years of experience.

Following these findings, we intend to perform more validations, standardizing the clinical cases to evaluate and their evaluation. The reflection condition is a strategy described by experts in diagnostic reasoning [12, 13, 14] in which diagnoses follow a standardized, structured procedure to arrive at the most likely differential diagnosis for a specific case. This approach could yield better intra-evaluator variability in future studies. For now, we intend to perform this validation in three specific subgroups (students, general physicians, specialists), using a bigger sample size to draw valid conclusions for each.

## Data Availability

All data produced in the present study are available upon reasonable request to the authors

## Notes

### Competing Interest Statement

All authors declare that they work at Arkangel AI

### Clinical Trial

According to the checklist from clinicaltrials.gov https://docsend.com/view/d2aqbx964k6uni92 our study is not an applicable clinical trial. It is interventional however it is not in US territory, under and FDA drug or interventional device application, does not involve a drug, biological or device product manufactured or regulated by the FDA.

### Clinical Protocols

https://cdn.clinicaltrials.gov/documents/ACT_Checklist.pdf

### Funding Statement

This study did not receive any funding

### Author Declarations

All of the participants of the study signed an informed consent form to use the data collected from the quiz to make this study available at this link https://docsend.com/view/d2aqbx964k6uni92.

## References

[1] Gilson, A., Safranek, C.W., Huang, T., Socrates, V., Chi, L., Taylor, R.A., et al., How Does ChatGPT Perform on the United States Medical Licensing Examination (USMLE)? The Implications of Large Language Models for Medical Education and Knowledge Assessment, JMIR Med Educ, 2023;9:e45312.

[2] Thirunavukarasu, A.J., Ting, D.S.J., Elangovan, K., Gutierrez, L., Tan, T.F., Ting, D.S.W., Large language models in medicine, Nat Med, 2023;29(8):1930–40.

[3] Singhal, K., Tu, T., Gottweis, J., Sayres, R., Wulczyn, E., Hou, L., et al., Towards Expert-Level Medical Question Answering with Large Language Models, arXiv preprint, 2023.

[4] Qin, Z., Luo, C., Wang, Z., Jiang, H., Sun, Y., Relational Database Augmented Large Language Model, arXiv preprint, 2024.

[5] Llano, I., Villa, M.C., Castano-Villegas, N., Martinez, J., Guevara, M.F., Zea, J., et al., Medsearch: A Conversational Agent for Real-Time, Evidence-Based Medical Question-Answering, SSRN Electronic Journal, 2025.

[6] Jin, Q., Dhingra, B., Liu, Z., Cohen, W.W., Lu, X., PubMedQA: A Dataset for Biomedical Research Question Answering, arXiv preprint, 2019.

[7] Labrak, Y., Bazoge, A., Dufour, R., Rouvier, M., Morin, E., Daille, B., et al., FrenchMedMCQA: A French Multiple-Choice Question Answering Dataset for Medical domain, arXiv preprint, 2023.

[8] Kotschenreuther, K., EHR-DS-QA: A Synthetic QA Dataset Derived from Medical Discharge Summaries for Enhanced Medical Information Retrieval Systems, PhysioNet, 2024.

[9] Pal, A., Umapathi, L.K., Sankarasubbu, M., MedMCQA: A Large-scale Multi-Subject Multi-Choice Dataset for Medical domain Question Answering, Proceedings of the Conference on Health, Inference, and Learning, PMLR, 2022:248–60.

[10] Papers with Code, MedQA Benchmark (Question Answering), 2024.

[11] Sherani, A.M.K., Khan, M., Qayyum, M.U., Hussain, H.K., Synergizing AI and Healthcare: Pioneering Advances in Cancer Medicine for Personalized Treatment, Int J Multidiscip Sci Arts, 2024;3(2):270–7.

[12] Goh, E., Gallo, R., Hom, J., Strong, E., Weng, Y., Kerman, H., et al., Influence of a Large Language Model on Diagnostic Reasoning: A Randomized Clinical Vignette Study, medRxiv, 2024.

[13] Berner, E.S., Webster, G.D., Shugerman, A.A., Jackson, J.R., Algina, J., Baker, A.L., et al., Performance of Four Computer-Based Diagnostic Systems, N Engl J Med, 1994;330(25):1792–6.

[14] van der Lee, C., Gatt, A., van Miltenburg, E., Wubben, S., Krahmer, E., Best practices for the human evaluation of automatically generated text, Proceedings of the 12th International Conference on Natural Language Generation, Association for Computational Linguistics, 2019:355–68.

[15] Abeysinghe, B., Circi, R., The Challenges of Evaluating LLM Applications: An Analysis of Automated, Human, and LLM-Based Approaches, arXiv preprint, 2024.

[16] Ibrahim, L., Huang, S., Ahmad, L., Anderljung, M., Beyond static AI evaluations: advancing human interaction evaluations for LLM harms and risks, arXiv preprint, 2024.

[17] Badshah, S., Sajjad, H., Reference-Guided Verdict: LLMs-as-Judges in Automatic Evaluation of Free-Form Text, arXiv preprint, 2024.

[18] Hamzic, D., Wurzenberger, M., Skopik, F., Landauer, M., Rauber, A., Evaluation and Comparison of Open-Source LLMs Using Natural Language Generation Quality Metrics, IEEE International Conference on Big Data, 2024:5342–51.

[19] Confident AI, LLM Evaluation Metrics: The Ultimate LLM Evaluation Guide, 2024.

[20] Roychowdhury, S., Soman, S., Ranjani, H.G., Gunda, N., Chhabra, V., Bala, S.K., Evaluation of RAG Metrics for Question Answering in the Telecom Domain, arXiv preprint, 2024.

[21] Mapping the Future of Healthcare: Six Practical Foresight Methods You Can Use Today [Internet]. The Medical Futurist. 2025 [cited 2025 Apr 30]. Available from: https://medicalfuturist.com/mapping-the-future-of-healthcare-six-practical-foresight-methods-you-can-use-today/

