## Supplementary Materials for "Real-world Validation of MedSearch: a conversational agent for real-time, evidence-based medical question-answering"

**SUPPLEMENTARY MATERIAL**

**Supplementary Material 1. Informed consent form signed by participants of the study (original version in Spanish) English translation obtained with ChatGPT.**

**I**NFORMED CONSENT FOR THE STUDY:

Project Title:
 Validation study of a conversational agent
 (MedSearch, first version called Vitruvius) for real-time medical question answering
 VITRUVIUS: A conversational agent for real-time evidence-based medical question answering
 Principal Investigator: Natalia Castaño Villegas

I, ______________________, identified with ID __________________________,
 declare that:
 • I have understood the written and/or verbal information provided to me.
 • I have been able and am able to ask any questions I deem necessary about the study.
 • My participation is voluntary.
 • I allow the recording of my screen during the meeting, for the purpose of evidence registration for the research (if applicable).
 • I have been informed about:

1. The objectives of the study and its procedures.
2. The benefits and inconveniences of the process.
3. The procedure and purpose for which my personal data will be used and the guarantees of compliance with current legal regulations.
4. That I can revoke my consent at any time (without needing to explain the reason) and request the deletion of my personal data.
5. That I have the right to access and rectify my personal data.

• I authorize Arkangel AI or its designees to use the information I provide for analytical purposes, to prepare studies and publish them.
 • Arkangel AI is not violating any confidentiality agreements, personal copyrights, or intellectual property rights by using the information provided, regardless of whether the studies result in any publication or commercial product.

I CONSENT TO PARTICIPATE IN THIS STUDY

(Write YES or NO)

To confirm all of the above, I sign below:

Date: _____________________
 Signature: _____________________

Investigator's Name: Natalia Castaño Villegas
 Investigator's Signature:

SECTION FOR REVOCATION OF CONSENT

I,
 ……………………………………………………………………………………………
 Revoke my consent to participate in the process signed above.

Signature and Date of Revocation

**Supplementary Material 2. Source list or prompt per workflow.**

| **Workflow** | **Sources/Prompt** |
| --- | --- |
| Clinical | "https://pubmed.ncbi.nlm.nih.gov/",  "https://scholar.google.com/",  "https://www.cochranelibrary.com/",  "https://clinicaltrials.gov/",  "https://www.embase.com/",  "https://www.sciencedirect.com/",  "https://jamanetwork.com/",  "https://www.bmj.com/",  "https://journals.plos.org/plosmedicine/",  "https://onlinelibrary.wiley.com/",  "https://www.nih.gov/",  "https://www.scopus.com/",  "https://www.uptodate.com/",  "https://www.webofscience.com/",  "https://eric.ed.gov/",  "https://www.who.int/hinari/en/",  "https://www.medrxiv.org/",  "https://www.biomedcentral.com/",  "https://www.researchgate.net/",  "https://www.scielo.org/",  "https://www.semanticscholar.org/",  "https://portal.fiocruz.br/",  "https://www.insp.mx/",  "https://www.ins.gov.co/",  "https://www.isciii.es/",  "https://www.canada.ca/en/services/health.html",  "https://www.nhmrc.gov.au/",  "https://www.argentina.gob.ar/salud/",  "https://www.gov.uk/government/organisations/department-of-health-and-social-care",  "https://www.aihw.gov.au/",  "https://www.santepubliquefrance.fr/" |
| Pharma | "https://dailymed.nlm.nih.gov/",  "https://www.drugs.com/"; |
| Differential | Performs a search based on the following prompt objective: "As a medical expert, generate a comprehensive clinical guideline and publications search given the patient's symptoms and medical history to determine that will support the differential diagnosis" |
| Clinical Plan | Performs a search based on the following prompt objective: "As a medical professional, conduct a comprehensive search supporting clinical guidelines and publications that will support a clinical plan for the given query" |
| Writer | Does not search. Designed for requests that only require writing support. |
| Pharmaceutical Industry | https://www.statista.com  https://www.researchandmarkets.com  https://www.marketwatch.com  https://www.fda.gov/  https://www.ema.europa.eu/  https://www.picscheme.org/ |

****Sources are updated frequently based on real-time feedback from users. The sources above correspond to those included at the time of the study.***

**Supplementary Material 3.**

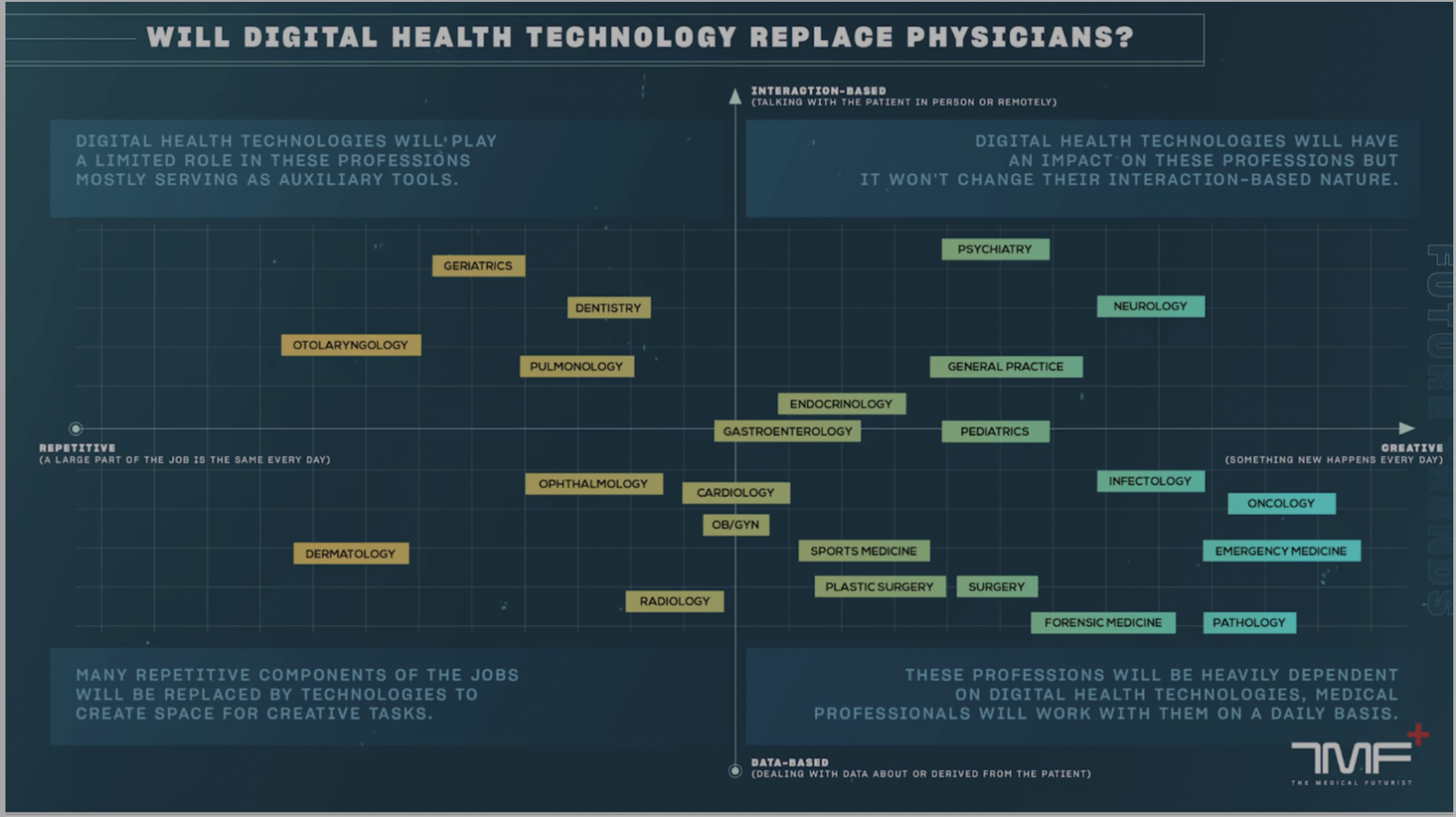

**Supplementary Material 4. Average scores for each of the six validity questions for each group per specialty.**

| **Specialty / Evaluation Question** | | **Pediatrics** | **Orthopedics** | **Psychiatry** | **Gynecology** |
| --- | --- | --- | --- | --- | --- |
| **Response Accuracy**  **(mean, 95% CI)** | **Group A** | 2.76  (2.69, 2.84) | 2.81  (2.74, 2.88) | 2.56  (2.47, 2.64) | 2.61  (2.53, 2.69) |
|  | **Group B** | 2.38  (2.26, 2.50) | 2.49  (2.39, 2.60) | 2.20  (2.10, 2.29) | 2.34  (2.24, 2.43) |
|  | **Mann Whitney's U** | <0.05 | <0.05 | <0.05 | <0.05 |
| **Medical Consensus**  **(mean, 95% CI)** | **Group A** | 2.91  (2.87, 2.95) | 2.47  (2.35, 2.59) | 2.98  (2.96, 3.00) | 2.88  (2.83, 2.93) |
|  | **Group B** | 2.72  (2.65, 2.80) | 2.42  (2.30, 2.55) | 2.93  (2.90, 2.97) | 2.78  (2.71, 2.85) |
|  | **Mann Whitney's U** | <0.05 | 0.51 | <0.05 | <0.05 |
| **Demographic Bias**  **(mean, 95% CI)** | **Group A** | 2.75  (2.67, 2.84) | 2.99  (2.97, 3.00) | 2.99  (2.97, 3.00) | 2.68  (2.61, 2.76) |
|  | **Group B** | 2.49  (2.37, 2.60) | 2.98  (2.95, 3.00) | 2.99  (2.98, 3.00) | 2.30  (2.19, 2.42) |
|  | **Mann Whitney's U** | <0.05 | 0.26 | 0.93 | <0.05 |
| **Treatment Bias**  **(mean, 95% CI)** | **Group A** | 2.88  (2.83, 2.94) | 2.95  (2.91, 2.99) | 3.00  (3.00, 3.00) | 2.73  (2.65, 2.81) |
|  | **Group B** | 2.67  (2.57, 2.77) | 2.88  (2.81, 2.94) | 3.00  (3.00, 3.00) | 2.37  (2.26, 2.48) |
|  | **Mann Whitney's U** | <0.05 | 0.05 | - | <0.05 |
| **Updated Information**  **(mean, 95% CI)** | **Group A** | 2.91  (2.86, 2.96) | 3.00  (3.00, 3.00) | 3.00  (2.99, 3.00) | 2.67  (2.59, 2.75) |
|  | **Group B** | 2.73  (2.64, 2.82) | 3.00  (3.00, 3.00) | 2.99  (2.98, 3.00) | 2.29  (2.17, 2.40) |
|  | **Mann Whitney's U** | <0.05 | - | 0.49 | <0.05 |
| **Patient Risk**  **(mean, 95% CI)** | **Group A** | 2.89  (2.83, 2.94) | 2.97  (2.94, 3.00) | 2.98  (2.96, 3.00) | 2.75  (2.68, 2.82) |
|  | **Group B** | 2.73  (2.64, 2.82) | 2.94  (2.89, 2.98) | 2.94  (2.90, 2.97) | 2.40  (2.29, 2.51) |
|  | **Mann Whitney's U** | <0.05 | 0.27 | <0.05 | <0.05 |

**Supplementary Material 5. Average scores for each of the six validity questions for each group per question type.**

| **Evaluation Question /**  **Question Type** | | **Diagnostic** | **Management** | **Research** | **General Knowledge** |
| --- | --- | --- | --- | --- | --- |
| **Response Accuracy**  **(mean, 95% CI)** | **Group A** | 2.76  (2.69, 2.83) | 2.61  (2.53, 2.69) | 2.73  (2.66, 2.80) | 2.63  (2.55, 2.72) |
|  | **Group B** | 2.41  (2.30, 2.51) | 2.27  (2.17, 2.38) | 2.37  (2.27, 2.48) | 2.36  (2.25, 2.46) |
|  | **Mann Whitney's U** | <0.05 | <0.05 | <0.05 | <0.05 |
| **Medical Consensus**  **(mean, 95% CI)** | **Group A** | 2.95  (2.93, 2.98) | 2.90  (2.86, 2.94) | 2.96  (2.94, 2.99) | 2.42  (2.30, 2.54) |
|  | **Group B** | 2.84  (2.78, 2.89) | 2.84  (2.78, 2.90) | 2.80  (2.73, 2.86) | 2.38  (2.26, 2.51) |
|  | **Mann Whitney's U** | <0.05 | 0.225 | <0.05 | 0.58 |
| **Demographic Bias**  **(mean, 95% CI)** | **Group A** | 2.93  (2.88, 2.97) | 2.79  (2.72, 2.86) | 2.84  (2.78, 2.90) | 2.86  (2.80, 2.92) |
|  | **Group B** | 2.72  (2.63, 2.82) | 2.64  (2.55, 2.73) | 2.61  (2.50, 2.72) | 2.80  (2.72, 2.87) |
|  | **Mann Whitney's U** | <0.05 | <0.05 | <0.05 | 0.05 |
| **Treatment Bias**  **(mean, 95% CI)** | **Group A** | 2.91  (2.86, 2.96) | 2.81  (2.74, 2.87) | 2.92  (2.87, 2.97) | 2.92  (2.87, 2.97) |
|  | **Group B** | 2.73  (2.64, 2.82) | 2.63  (2.54, 2.73) | 2.71  (2.62, 2.80) | 2.83  (2.76, 2.90) |
|  | **Mann Whitney's U** | <0.05 | <0.05 | <0.05 | <0.05 |
| **Updated Information**  **(mean, 95% CI)** | **Group A** | 2.95  (2.92, 2.99) | 2.80  (2.74, 2.87) | 2.90  (2.85, 2.95) | 2.92  (2.87, 2.96) |
|  | **Group B** | 2.85  (2.78, 2.91) | 2.61  (2.50, 2.71) | 2.68  (2.59, 2.77) | 2.88  (2.81, 2.93) |
|  | **Mann Whitney's U** | <0.05 | <0.05 | <0.05 | 0.14 |
| **Patient Risk**  **(mean, 95% CI)** | **Group A** | 2.90  (2.85, 2.95) | 2.82  (2.75, 2.88) | 2.93  (2.90, 2.97) | 2.93  (2.89, 2.97) |
|  | **Group B** | 2.78  (2.70, 2.86) | 2.63  (2.53, 2.72) | 2.71  (2.62, 2.80) | 2.89  (2.83, 2.94) |
|  | **Mann Whitney's U** | <0.05 | <0.05 | <0.05 | 0.14 |
